# Association between Insurance Type and Extended Length of Stay in Urban & Rural Vermont Hospitals

**DOI:** 10.1101/2024.06.23.24309359

**Authors:** Patrick C. Payne, Mia Klonsky, Katrina Moreau, Alyssa Oviedo, Sarah Nowak

## Abstract

**Purpose:** To evaluate the link between insurance status and patient length of stay (LOS) for inpatient admissions in rural and urban hospitals in Vermont.

**Methods:** We conducted a cross-sectional study utilizing 2016 to 2020 data from the Vermont Uniform Hospital at Data Discharge System (VUHDDS). Vermont residents 18-64 years of age admitted for heart and circulatory illnesses who spent least one day as inpatient at one of Vermont’s 14 hospitals were included. Frequency statistics were run to determine distribution of sample characteristics and a two-side Z-test was conducted to compare differences between normal and extended lengths of stay. Three multivariate logistic regression models were utilized to control for confounding factors and identify differences in lengths of stay and mortality.

**Findings:** Private insurance was more common among patients with a normal LOS (46%) and public insurance more common among patients with extended lengths of stay (54%). Rural Medicare patients have 0.77 (CI: 0.66-0.90) times lower odds of extended LOS, which is distinct from the 95% confidence interval for urban patients (CI: 0.94-1.46). Urban Medicaid insured patients had 1.413 (CI: 1.15-1.74) times greater odds of an extended LOS, which is outside of the 95% confidence interval for rural patients (CI: 0.80-1.09).

**Conclusions:** In conclusion, the rurality of a patient’s residence appears to impact health outcomes for cardiac related discharges for individuals in Vermont related to their insurer. Further studies with more demographic data are needed to better understand the implications of these findings.

## Introduction

In the United States, a patient’s insurance status – private, public, or no health insurance - often dictates many aspects of the care they can receive. The disparities between public and private insurance are noteworthy, with publicly insured individuals having significantly higher mortality rates for common conditions and less access to medical care. ^1,2^ In addition to inequities based on insurance type, the difference between care in rural and urban settings makes this gap even larger. Rural areas often see lower private health insurance rates among their populations. ^3^ Rural areas tend to have much higher rates of Medicaid insurance than their urban counterparts in patient populations under 65. This issue, coupled with fewer options for care, can lead to higher mortality rates for diseases like cancer and lower life expectancy within the population. ^4,5^ Studies have found many of these rural communities that lack high rates of both public and private insurance coverage have a low understanding of health insurance and care options, causing the barriers to care to be more significant. ^6–8^

Many studies have shown place indicates whether a person is insured, and rural communities tend to have much lower rates than urban ones. ^9,10^ There is also substantial evidence that rural communities have poorer health outcomes than patients in urban communities. ^11,12^ By researching insurance type compared to patient outcomes such as length of stay (LOS), discharge status, and diagnosis in Vermont, we intend to better understand the role health insurance plays in patient care between rural and urban in-patient environments for patients aged 18 and 64 years. This investigation will contribute to the growing body of research supporting health insurance reform by filling the gaps in our understanding of health insurance’s health benefits as it relates to patient outcomes in rural and urban settings.

## Methods

We conducted a cross-sectional study utilizing 2016 to 2020 data from the Vermont Uniform Hospital Data Discharge System (VUHDDS). The VUHDDS database is managed by the Vermont Department of Health and administered by the Green Mountain Care Board. This database contains emergency department and inpatient data for all 14 acute care hospitals in Vermont and is available as a limited use Data Set which includes reasons for admission, treatments, charges, diagnoses, and discharge data. This database was selected as Vermont was an early adopter of Medicaid expansion and has documented efforts of payer reform dating back to 2009. ^13,14^

### Study Population

The study population included all Vermont residents 18-64 years of age who spent at least one day as inpatient at one of Vermont’s 14 hospitals between 2016 and 2020. Sex and age were the only socio-demographic variables available in the database and both were included. The hospital admission data was limited to patients who were transferred from a hospital, non-health care facility point of origin, and clinical or doctor office referral. The hospital discharge data utilized were patients transferred to another acute hospital, skilled nursing facility, home (own or family care), home health, departure against medical advice, and death. Case mix index (CMI) is determined based on the Medicare Severity Diagnosis Related Groups associated with each patient and the severity and complexity of their disease. The CMI was used as the average of relative weights for the patients assigned to a specific population or group.

### Exclusion Criteria

Individuals were excluded if they were not a resident of Vermont, were <18 or >64 years of age, and did not have a major diagnostic category of a “heart and circulatory” diagnosis. Since we were only interested in insurance coverage by type, we excluded patients with worker’s compensation or no charge for coverage of payment.

### Statistical Analysis

SPSS version 26 was utilized to clean the data and for all computational methods. Frequency statistics were run to determine the distribution of sex, age groups, admission source and type, discharge status and year, hospital service area and rurality, and principle payer (insurance type). A two-sided Z-test was run to determine the difference between frequency statistics for normal and extended LOS. Additionally, individuals were coded as a rural or urban Vermont resident by hospital service area (HSA). Vermont has 14 hospitals with the Burlington hospital coded as urban and the other 13 coded as rural based off the U.S. Office of Management and Budget definition or rurality for the location of the hospital in the HSA. Frequency statistics were also run to determine the variability in insurance status (exposure) using the principal payment source data. We then performed descriptive analysis on LOS, producing Table 1.

**TABLE 1.** Sample Characteristics for patient samples having an extended and normal LOS. Statistically significant differences between the two categories are indicated a p-value less than 0.05. No test of comparison is performed for continuous variables.

|  | Normal LOS<br>(N=6137) | Extended LOS<br>(N=1629) | P value |
| --- | --- | --- | --- |
| **Principal Payer** |  |  |  |
| Medicare | 1349 (21.98%) | 521 (31.98%) | <0.05 |
| Medicaid | 1470 (23.95%) | 446 (27.38%) | <0.05 |
| CHAMPUS | 65 (1.06%) | 20 (1.23%) |  |
| Other Government | 42 (0.68%) | 11 (0.68%) |  |
| Self Pay | 195 (3.18%) | 36 (2.21%) | <0.05 |
| MVP | 91 (1.48%) | 19 (1.17%) |  |
| Commercial Insurance | 753 (12.27%) | 178 (10.93%) |  |
| HMO | 248 (4.04%) | 66 (4.05%) |  |
| BCBSVT | 1924 (31.35%) | 332 (20.38%) | <0.05 |
| **Age Group** |  |  |  |
| 18-24 | 69 (1.12%) | 18 (1.10%) |  |
| 25-29 | 60 (0.98%) | 25 (1.53%) |  |
| 30-34 | 104 (1.69%) | 34 (2.09%) |  |
| 35-39 | 192 (3.13%) | 52 (3.19%) |  |
| 40-44 | 319 (5.20%) | 66 (4.05%) |  |
| 45-49 | 577 (9.40%) | 121 (7.43%) | <0.05 |
| 50-54 | 1044 (17.01%) | 273 (16.76%) |  |
| 55-59 | 1634 (26.63%) | 408 (25.05%) |  |
| 60-64 | 2138 (34.84%) | 632 (38.80%) | <0.05 |
| **Admission Source** |  |  |  |
| Transfer from a Hospital | 1261 (20.55%) | 328 (20.14%) |  |
| Clinic or Doctor Office Referral | 295 (4.81%) | 93 (5.71%) |  |
| Non-Health Care Facility Point of Origin | 4581 (74.65%) | 1208 (74.16%) |  |
| **Admission Type** |  |  |  |
| Urgent | 1299 (21.17%) | 359 (22.04%) |  |
| Elective | 638 (10.40%) | 215 (13.20%) | <0.05 |
| Emergency | 4200 (68.44%) | 1055 (64.76%) | <0.05 |
| **Discharge Status** |  |  |  |
| To Another Acute Hospital | 710 (11.57%) | 75 (4.60%) | <0.05 |
| To a Skilled Nursing Facility | 58 (0.95%) | 149 (9.15%) | <0.05 |
| Home - Home Health | 671 (10.93%) | 742 (45.55%) | <0.05 |
| Against Medical Advice | 113 (1.84%) | 39 (2.39%) |  |
| Died | 83 (1.35%) | 31 (1.90%) |  |
| Home - Own or Family Care | 4502 (73.36%) | 593 (36.40%) | <0.05 |

**TABLE 1 cont.** Sample Characteristics for patient samples having an extended and normal LOS. Statistically significant differences between the two categories are indicated a p-value less than 0.05. No test of comparison is performed for continuous variables.
|  | Normal LOS<br>(N=6137) | Extended LOS<br>(N=1629) | P value |
| --- | --- | --- | --- |
| **Year of Discharge** |  |  |  |
| 2016 | 1186 (19.33%) | 302 (18.54%) |  |
| 2017 | 1242 (20.24%) | 318 (19.52%) |  |
| 2018 | 1283 (20.91%) | 330 (20.26%) |  |
| 2019 | 1287 (20.97%) | 349 (21.42%) |  |
| 2020 | 1139 (18.56%) | 330 (20.26%) |  |
| **Sex** |  |  |  |
| Female | 2197 (35.80%) | 637 (39.10%) | <0.05 |
| Male | 3940 (64.20%) | 992 (60.90%) | <0.05 |
| **Hospital Service Area** |  |  |  |
| Burlington | 1875 (30.55%) | 472 (28.97%) |  |
| Morrisville | 383 (6.24%) | 87 (5.34%) |  |
| Randolph | 147 (2.40%) | 41 (2.52%) |  |
| Newport | 171 (2.79%) | 45 (2.76%) |  |
| St. Johnsbury | 159 (2.59%) | 37 (2.27%) |  |
| St. Albans | 725 (11.81%) | 200 (12.28%) |  |
| Middlebury | 282 (4.60%) | 90 (5.52%) |  |
| Rutland | 907 (14.78%) | 236 (14.49%) |  |
| Bennington | 222 (3.62%) | 47 (2.89%) |  |
| Springfield | 147 (2.40%) | 42 (2.58%) |  |
| White River Jct. | 67 (1.09%) | 12 (0.74%) |  |
| Brattleboro | 110 (1.79%) | 32 (1.96%) |  |
| Barre | 942 (15.35%) | 288 (17.68%) | <0.05 |
| **Is the patient from a rural or urban area?** |  |  |  |
| Urban | 1875 (30.55%) | 472 (28.97%) |  |
| Rural | 4262 (69.45%) | 1157 (71.03%) |  |
| Mean charge per day (95% CI) | \$10,546 (10342 , 10755) | \$6,442 (6273, 6612) | |
| Median Number of days in a specialty care unit | 0 | 0 |  |
| Mean Case Mix Index Weight (95% CI) | 1.593 (1.561, 1.626) | 2.985 (2.869, 3.100) |  |

CI = Confidence Interval

Our outcome measure, LOS, was analyzed using multivariate logistic regression with three regressions models being designed, one on unstratified and the others with data stratified by rurality of the patient. The strata that were formed are urban patients and rural patients. LOS was converted to binary variables by defining the quartiles for the variable and identifying in the upper quartile for the LOS at the patient’s hospital with the the same major diagnostic category, consistent with other publications. ^15,16^ Results from the multivariate logistic regression analyses are reported for the study period with adjusted odds ratios and 95% confidence intervals. We completed a stepwise regression analysis to select variables for inclusion that were not initially identified through the purposeful selection of covariates. Insurance status was analyzed by principal payer type, rurality by hospital service area, and mortality by discharge status.

The extended LOS regression model contained admission type, admission source, age group, biological sex, principal payer, year of discharge, hospital service area, patient charges divided by LOS, and number of specialty care days. This model included discharge status as a covariate to account for censorship. Analysis of missingness was conducted to determine if missing values had any correlations. Based on the missingness findings, only complete entries were utilized in the analysis. All tests were two-sided with a level of significance of 0.05. The University of Vermont Institutional Review Board has reviewed this project and determined that it qualifies as exempt from additional review.

## Results

The sample contained 7,766 discharges of patients after applying the exclusion and inclusion criteria (Table 1). The sample consisted of 6,137 discharged patients that had a normal LOS and 1,629 discharged patients that had an extended LOS at the hospital where they received treatment. The patients were mostly male (63.5%), aged 60-64 (35.7%) and from a rural hospital service area (69.8%). The most common public insurance was Medicaid (24.7%) and most common private insurance BlueCross BlueShield of Vermont (BCBSVT) (29.0%).

The patients that experienced an extended LOS showed higher case complexities, with an average case mix index of 2.985. Having BCBSVT, a private insurer, was significantly more common among those who experienced a normal LOS (31.35%) and Medicare & Medicaid were significantly more common among those with an extended LOS (31.98% & 27.38%). The average age of the extended LOS patients is marginally greater (54.7 years, 54.0 years). Patients with extended lengths of stay were mainly discharged to home health (45.55%), whereas normal LOS patients were discharged to their own or family care (73.36%).

After controlling for significant confounding factors, the multivariate logistic regression showed that patients with Commercial Insurance, CHAMPUS, or HMO insurance plans had odds of an extended LOS that were 1.32 (95% CI: 1.15 – 1.52), 2.035 (95% CI: 1.36-3.06), and 1.72 (95% CI: 1.40-2.12) times that of BCBSVT insured patients. The unstratified sample (Table 2) suggests that we are not able to conclude that patients with the Medicare and Medicaid have differences in LOS compared to the largest private insurer in Vermont, BCBSVT. However, stratifying by the patient being from a rural or urban hospital service area reveals a difference in the strata.

**TABLE 2.** Results of a multivariate logistic regression of extended LOS among patients discharged between 2016 and 2020. The data used in this model represents the entire dataset after inclusions and exclusion, i.e. the data is unstratified.

|  | OR | Lower CI | Upper CI | P value |
| --- | --- | --- | --- | --- |
| **Principal Payer** |  |  |  |  |
| Medicare | 0.934 | 0.825 | 1.058 | 0.283 |
| Medicaid | 1.123 | 0.995 | 1.268 | 0.060 |
| CHAMPUS | 2.035 | 1.356 | 3.055 | 0.001 |
| Other Government | 1.190 | 0.729 | 1.942 | 0.486 |
| Self Pay | 0.980 | 0.736 | 1.306 | 0.892 |
| MVP | 1.101 | 0.768 | 1.579 | 0.601 |
| Commercial Insurance | 1.321 | 1.146 | 1.522 | < 0.001 |
| HMO | 1.722 | 1.397 | 2.122 | < 0.001 |
| BCBSVT | REF | REF | REF |  |
| **Age Group** |  |  |  |  |
| 18-24 | 1.822 | 1.204 | 2.759 | 0.005 |
| 25-29 | 2.149 | 1.487 | 3.105 | < 0.001 |
| 30-34 | 0.995 | 0.727 | 1.361 | 0.973 |
| 35-39 | 1.380 | 1.081 | 1.761 | 0.010 |
| 40-44 | 1.021 | 0.830 | 1.257 | 0.845 |
| 45-49 | 0.941 | 0.797 | 1.112 | 0.474 |
| 50-54 | 1.144 | 1.008 | 1.297 | 0.036 |
| 55-59 | 1.013 | 0.908 | 1.130 | 0.818 |
| 60-64 | REF | REF | REF |  |
| **Admission Source** |  |  |  |  |
| Transfer from a Hospital | 1.803 | 1.553 | 2.093 | < 0.001 |
| Clinic or Doctor Office Referral | 1.004 | 0.777 | 1.296 | 0.977 |
| Non-Health Care Facility Point of Origin | REF | REF | REF |  |
| **Admission Type** |  |  |  |  |
| Urgent | 1.267 | 1.099 | 1.459 | 0.001 |
| Elective | 0.820 | 0.726 | 0.927 | 0.001 |
| Emergency | REF | REF | REF |  |
| **Discharge Status** |  |  |  |  |
| To Another Acute Hospital | 0.644 | 0.518 | 0.800 | < 0.001 |
| To a Skilled Nursing Facility | 19.704 | 14.987 | 25.906 | < 0.001 |
| Home - Home Health | 8.293 | 7.515 | 9.152 | < 0.001 |
| Against Medical Advice | 2.136 | 1.573 | 2.900 | < 0.001 |
| Died | 4.487 | 3.392 | 5.936 | < 0.001 |
| Home - Own or Family Care | REF | REF | REF |  |

OR = odds ratio, CI = Confidence Interval

**TABLE 2 cont.** Results of a multivariate logistic regression of extended LOS among patients discharged between 2016 and 2020. The data used in this model represents the entire dataset after inclusions and exclusion, i.e. the data is unstratified.
|  | OR | Lower CI | Upper CI | P value |
| --- | --- | --- | --- | --- |
| **Year of Discharge** |  |  |  |  |
| 2016 | REF | REF | REF |  |
| 2017 | 0.984 | 0.860 | 1.127 | 0.820 |
| 2018 | 1.130 | 0.985 | 1.296 | 0.080 |
| 2019 | 1.271 | 1.108 | 1.457 | 0.001 |
| 2020 | 1.725 | 1.499 | 1.985 | < 0.001 |
| **Sex** |  |  |  |  |
| Female | 0.834 | 0.761 | 0.914 | < 0.001 |
| Male | REF | REF | REF |  |
| **Hospital Service Area** |  |  |  |  |
| Burlington | 1.339 | 1.168 | 1.535 | < 0.001 |
| Morrisville | 0.813 | 0.666 | 0.993 | 0.043 |
| Randolph | 1.192 | 0.822 | 1.727 | 0.354 |
| Newport | 0.815 | 0.598 | 1.110 | 0.194 |
| St. Johnsbury | 0.835 | 0.584 | 1.194 | 0.324 |
| St. Albans | 1.152 | 0.981 | 1.353 | 0.085 |
| Middlebury | 1.244 | 1.016 | 1.522 | 0.034 |
| Rutland | 0.974 | 0.829 | 1.144 | 0.748 |
| Bennington | 0.253 | 0.178 | 0.360 | < 0.001 |
| Springfield | 0.609 | 0.418 | 0.887 | 0.010 |
| White River Jct. | 0.870 | 0.505 | 1.500 | 0.617 |
| Brattleboro | 0.813 | 0.533 | 1.240 | 0.336 |
| Barre | REF | REF | REF |  |
| Log of the mean charge per day | 0.048 | 0.040 | 0.057 | < 0.001 |
| Number of days in a specialty care unit | 0.984 | 0.980 | 0.989 | < 0.001 |

OR = odds ratio, CI = Confidence Interval

The strata show differences in LOS for rural and urban patients depending on the patient’s insurer. Notably, the large public insurers decrease the odds of an extended LOS in rural patients, but increase the odds of an extended LOS for urban patients (Table 3). Rural medicare patients have 0.781 (95% CI: 0.669-0.912) times lower odds of extended LOS, which is significantly different from the 95% confidence interval for urban patients (95% CI: 0.944-1.460). Urban medicaid insured patients had 1.413 (95% CI: 1.151-1.735) times greater odds of an extended LOS. The 95% confidence intervals for urban and rural medicaid insured patients are non-overlapping, which is distinct from the 95% confidence interval for rural patients (Urban 1.151-1.735, Rural: 0.798-1.086).

**TABLE 3.** Results from a stratified multivariate logistic regression. The strata represent the patients from urban hospital service areas and the rural hospital service areas. The model for the strata differ on one variable. The hospital service area variable is not included in the urban regression as there is only one area, Burlington.

|  | Rural Patient Stratum (n = 5419) |  |  |  | Urban Patient Stratum (n = 2347) |  |  |  |
| --- | --- | --- | --- | --- | --- | --- | --- | --- |
|  | OR | Lower CI | Upper CI | P value | OR | Lower CI | Upper CI | P value |
| **Principal Payer** |  |  |  |  |  |  |  |  |
| Medicare | 0.781 | 0.669 | 0.912 | 0.002 | 1.174 | 0.944 | 1.460 | 0.148 |
| Medicaid | 0.931 | 0.798 | 1.086 | 0.365 | 1.413 | 1.151 | 1.735 | 0.001 |
| CHAMPUS | 1.286 | 0.731 | 2.262 | 0.383 | 4.470 | 2.404 | 8.309 | < 0.001 |
| Other Government | 0.946 | 0.501 | 1.788 | 0.865 | 1.157 | 0.507 | 2.644 | 0.729 |
| Self Pay | 0.720 | 0.493 | 1.052 | 0.090 | 1.519 | 0.955 | 2.419 | 0.078 |
| MVP | 1.227 | 0.751 | 2.004 | 0.413 | 0.832 | 0.486 | 1.426 | 0.503 |
| Commercial Insurance | 0.979 | 0.813 | 1.179 | 0.825 | 1.897 | 1.509 | 2.385 | < 0.001 |
| HMO | 1.396 | 1.039 | 1.876 | 0.027 | 2.508 | 1.849 | 3.402 | < 0.001 |
| BCBSVT | REF | REF | REF |  | REF | REF | REF |  |
| **Age Group** |  |  |  |  |  |  |  |  |
| 18-24 | 1.796 | 1.012 | 3.188 | 0.045 | 1.780 | 0.950 | 3.334 | 0.072 |
| 25-29 | 2.261 | 1.394 | 3.669 | 0.001 | 2.085 | 1.170 | 3.716 | 0.013 |
| 30-34 | 0.897 | 0.600 | 1.341 | 0.596 | 1.028 | 0.608 | 1.739 | 0.917 |
| 35-39 | 1.089 | 0.795 | 1.491 | 0.597 | 1.874 | 1.243 | 2.823 | 0.003 |
| 40-44 | 1.491 | 1.158 | 1.919 | 0.002 | 0.426 | 0.279 | 0.651 | < 0.001 |
| 45-49 | 0.919 | 0.743 | 1.137 | 0.438 | 1.043 | 0.785 | 1.386 | 0.772 |
| 50-54 | 1.081 | 0.921 | 1.267 | 0.340 | 1.224 | 0.991 | 1.511 | 0.061 |
| 55-59 | 0.904 | 0.787 | 1.039 | 0.154 | 1.217 | 1.015 | 1.460 | 0.034 |
| 60-64 | REF | REF | REF |  | REF | REF | REF |  |
| **Admission Source** |  |  |  |  |  |  |  |  |
| Transfer from a Hospital | 2.361 | 1.983 | 2.811 | < 0.001 | 1.045 | 0.697 | 1.568 | 0.830 |
| Clinic or Doctor Office Referral | 0.807 | 0.587 | 1.109 | 0.186 | 1.788 | 1.141 | 2.801 | 0.011 |
| Non-Health Care Facility Point of Origin | REF | REF | REF |  | REF | REF | REF |  |
| **Admission Type** |  |  |  |  |  |  |  |  |
| Urgent | 1.429 | 1.209 | 1.688 | < 0.001 | 1.079 | 0.799 | 1.455 | 0.621 |
| Elective | 1.434 | 1.212 | 1.697 | < 0.001 | 0.464 | 0.383 | 0.561 | < 0.001 |
| Emergency | REF | REF | REF |  | REF | REF | REF |  |
| **Discharge Status** |  |  |  |  |  |  |  |  |
| To Another Acute Hospital | 0.649 | 0.511 | 0.824 | < 0.001 | 1.502 | 0.796 | 2.832 | 0.209 |
| To a Skilled Nursing Facility | 17.660 | 12.820 | 24.326 | < 0.001 | 25.231 | 14.540 | 43.783 | < 0.001 |
| Home - Home Health | 8.135 | 7.166 | 9.234 | < 0.001 | 8.797 | 7.456 | 10.380 | < 0.001 |
| Against Medical Advice | 1.943 | 1.337 | 2.825 | < 0.001 | 3.499 | 1.931 | 6.341 | < 0.001 |
| Died | 3.876 | 2.686 | 5.593 | < 0.001 | 4.049 | 2.618 | 6.261 | < 0.001 |
| Home - Own or Family Care | REF | REF | REF |  | REF | REF | REF |  |

OR = odds ratio, CI = Confidence Interval

**TABLE 3 cont.** Results from a stratified multivariate logistic regression.
|  | Rural Patient Stratum (n = 5419) |  |  |  | Urban Patient Stratum (n = 2347) |  |  |  |
| --- | --- | --- | --- | --- | --- | --- | --- | --- |
|  | OR | Lower CI | Upper CI | P value | OR | Lower CI | Upper CI | P value |
| **Year of Discharge** |  |  |  |  |  |  |  |  |
| 2016 | REF | REF | REF |  | REF | REF | REF |  |
| 2017 | 0.982 | 0.831 | 1.162 | 0.837 | 1.001 | 0.789 | 1.269 | 0.995 |
| 2018 | 1.110 | 0.935 | 1.318 | 0.233 | 1.064 | 0.837 | 1.352 | 0.613 |
| 2019 | 1.234 | 1.040 | 1.464 | 0.016 | 1.321 | 1.040 | 1.679 | 0.022 |
| 2020 | 1.470 | 1.227 | 1.760 | < 0.001 | 2.370 | 1.871 | 3.002 | < 0.001 |
| **Sex** |  |  |  |  |  |  |  |  |
| Female | 0.925 | 0.824 | 1.039 | 0.188 | 0.664 | 0.566 | 0.779 | < 0.001 |
| Male | REF | REF | REF |  | REF | REF | REF |  |
| **Hospital Service Area** |  |  |  |  |  |  |  |  |
| Burlington | - | - | - | - | - | - | - | - |
| Morrisville | 1.266 | 0.865 | 1.852 | 0.014 | - | - | - | - |
| Randolph | 1.325 | 0.905 | 1.938 | 0.224 | - | - | - | - |
| Newport | 0.836 | 0.609 | 1.149 | 0.298 | - | - | - | - |
| St. Johnsbury | 0.964 | 0.663 | 1.401 | 0.665 | - | - | - | - |
| St. Albans | 1.141 | 0.966 | 1.347 | 0.235 | - | - | - | - |
| Middlebury | 1.326 | 1.079 | 1.631 | 0.019 | - | - | - | - |
| Rutland | 1.062 | 0.900 | 1.253 | 0.803 | - | - | - | - |
| Bennington | 0.317 | 0.223 | 0.451 | < 0.001 | - | - | - | - |
| Springfield | 0.741 | 0.507 | 1.084 | 0.016 | - | - | - | - |
| White River Jct. | 0.889 | 0.511 | 1.544 | 0.590 | - | - | - | - |
| Brattleboro | 0.951 | 0.615 | 1.469 | 0.635 | - | - | - | - |
| Barre | REF | REF | REF |  | - | - | - | - |
| Log of the mean charge per day | 0.025 | 0.020 | 0.032 | < 0.001 | 0.083 | 0.063 | 0.109 | < 0.001 |
| Number of days in a specialty care unit | 0.984 | 0.979 | 0.989 | < 0.001 | 0.986 | 0.974 | 0.998 | 0.022 |

OR = odds ratio, CI = Confidence Interval

There is a notable difference in the odds ratios for the rural patients with elective admissions and urban patients with elective admissions. Rural patients with elective admissions have 1.434 (95% CI: 1.212-1.697) times greater odds of an extended LOS. Whereas urban elective admissions have lower odds of extended LOS by a factor of 0.464 (95% CI: 0.383-0.561).

The models explained a moderate portion of the variability in LOS with Cox and Snell pseudo-R2 values of 0.301, 0.319, and 0.304 for the models unstratified data, rural patient stratum, and urban patient stratum as present in Tables 2 and 3. The F1-score for these models were 0.844, 0.844, and 0.849, respectively.

## Discussion

Focusing on the Vermont cardiac related discharges, we found that differences are present between the patient outcomes for individuals living in an urban population and those of the rural populations. The largest private insurer, BCBSVT, had the lowest odds of an extended LOS for urban patients. Being insured with Medicare was associated with lower odds of an extended LOS for rural patients. The differences in LOS were most notable in the reduction in odds of extended LOS for rural Medicare patients. This potentially arises from the higher proportion of publicly insured patients in rural settings. Although it is not fully understood, the tendency for rural individuals to receive less treatment could be due to smaller hospital size and a lower proportion of board-certified physicians. ^17^ This may also explain the higher odds of extended LOS for urban patients – they are receiving more treatments in a bigger hospital, with an intensive care unit and higher proportion of board-certified specialists. ^17,18^

The urban and rural populations also show a significant difference related to elective admissions, with urban patients receiving elective treatments having less than half the odds of an extended LOS and rural patients having higher odds when admitted for elective rather than emergency reasons. This may be related to the level of care received at the urban hospitals being higher, with elective surgeries and treatments being more routinely performed. Limitations of the study were due to the limited demographic data and clinical detail available for each patient.

Additional data regarding demographics such as socioeconomic status, race, and zip code were requested, but were unable to be obtained. Social determinants of health such as the socioeconomic status, race, and geographical location of a patient would have been beneficial to the analysis as the role these factors play in an individual’s health have been well documented and could have had an impact on patient’s LOS.

Many other studies analyzing the relationship between rural/urban healthcare settings and outcomes do so through one diagnostic category such as atrial fibrillation and myocardial infarction. ^19,20^ Many studies have not accounted for existing comorbidities or insurance status, the latter of which we believe can be a protective factor in healthcare settings. Additionally, studies like that of Davidson et al. show that uninsured patients have the shortest LOS while minimal difference exists between other insurance types ^21^ – however, this study does not address differences between rural and urban settings, which was important to address in this paper due to the disparities in health care between each location, as well as the rurality of Vermont.

This study concluded that there were no apparent differences in LOS dependent on insurance status, with Medicare, Medicaid, and BCBSVT patients experiencing similar odds of an extended LOS when unstratified. However, when adding the additional factor of rurality, the regression showed that rural patients had greater protection from an extended LOS when having a public insurer like Medicare and the opposite finding was present in urban patients. It is important to use this information to assess accessibility and impact of health insurance in Vermont (and other states like Vermont), as well as outcomes for patients dependent on whether they are rural or urban. Further research is needed that considers demographic data and utilizes stronger regression models to further analyze the trends of patient outcomes based on their insurance status and rural/urban location.

## Conclusion

The rurality of a patient appears to impact health outcomes for cardiac related discharges for patients in Vermont related to the patient’s insurer. Further studies with more demographic are needed to better understand the implications of these findings.

## Data Availability

Data can be accessed through the Green Mountain Care Board. Access to scripts used for data cleaning and processing are available upon request.

https://gmcboard.vermont.gov/form/vuhdds-puf

## Notes

### Competing Interest Statement

The authors have declared no competing interest.

### Funding Statement

This study did not receive any funding.

### Author Declarations

IRB of Univerisity of Vermont waived ethical approval for this work as an exept researh project. Study utilized deidentified dataset.

## References

[1] Woolhandler Steffie, Himmelstein David U. The relationship of health insurance and mortality: Is lack of insurance deadly? Ann. Intern. Med.. 2017;167:424–431.

[2] Hao Scarlett, Snyder Rebecca A, Irish William, Parikh Alexander A. Association of race and health insurance in treatment disparities of colon cancer: A retrospective analysis utilizing a national population database in the United States PLoS Med.. 2021;18:e1003842.

[3] Zahnd Whitney E, Murphy Cathryn, Knoll Marie, et al. The intersection of rural residence and minority race/ethnicity in cancer disparities in the United States Int. J. Environ. Res. Public Health. 2021;18:1384.

[4] Cross Sarah H, Califf Robert M, Warraich Haider J. Rural-urban disparity in mortality in the US from 1999 to 2019 JAMA. 2021;325:2312–2314.

[5] Iglehart John K. The challenging quest to improve rural health care N. Engl. J. Med.. 2018;378:473–479.

[6] Monnat Shannon M. The new destination disadvantage: Disparities in Hispanic health insurance coverage rates in metropolitan and nonmetropolitan new and established destinations Rural Sociol.. 2017;82:3–43.

[7] Edward Jean, Thompson Robin, Jaramillo Andrea. Availability of health insurance literacy resources fails to meet consumer needs in rural, Appalachian communities: Implications for state Medicaid waivers J. Rural Health. 2021;37:526–536.

[8] Erlangga Darius, Suhrcke Marc, Ali Shehzad, Bloor Karen. The impact of public health insurance on health care utilisation, financial protection and health status in low- and middle-income countries: A systematic review PLoS One. 2019;14:e0219731.

[9] Stone Lisa Cacari, Boursaw Blake, Bettez Sonia P, Larzelere Marley Tennille, Waitzkin Howard. Place as a predictor of health insurance coverage: A multivariate analysis of counties in the United States Health Place. 2015;34:207–214.

[10] Robinson Lara R, Holbrook Joseph R, Bitsko Rebecca H, et al. Differences in health care, family, and community factors associated with mental, behavioral, and developmental disorders among children aged 2-8 years in rural and urban areas - United States, 2011-2012 MMWR Surveill. Summ.. 2017;66:1–11.

[11] Andrilla C Holly A, Patterson Davis G, Garberson Lisa A, Coulthard Cynthia, Larson Eric H. Geographic variation in the supply of selected behavioral health providers Am. J. Prev. Med.. 2018;54:S199–S207.

[12] Morales Dawn A, Barksdale Crystal L, Beckel-Mitchener Andrea C. A call to action to address rural mental health disparities J. Clin. Transl. Sci.. 2020;4:463–467.

[13] Lin Yilu, Monnette Alisha, Shi Lizheng. Effects of medicaid expansion on poverty disparities in health insurance coverage International Journal for Equity in Health. 2021;20.

[14] Jones Craig, Watkins Lisa Dulsky, Samuelson Jenney, Morgan James, Price Terri, Hawkins Diane. Vermont Blueprint for Health: 2009 Annual Report 2010.

[15] Hoffman Ryan D, Danos Denise M, Lau Frank H. National health disparities in incisional hernia repair outcomes: An analysis of the Healthcare Cost and Utilization Project National Inpatient Sample (HCUP-NIS) 2012-2014 Surgery. 2021;169:1393–1399.

[16] Lingsma Hester F, Bottle Alex, Middleton Steve, Kievit Job, Steyerberg Ewout W, Mheen Perla J. Evaluation of hospital outcomes: the relation between length-of-stay, readmission, and mortality in a large international administrative database BMC Health Serv. Res.. 2018;18:116.

[17] Baldwin Laura-Mae, MacLehose Richard F, Hart L Gary, Beaver Shelli K, Every Nathan, Chan Leighton. Quality of care for acute myocardial infarction in rural and urban US hospitals J. Rural Health. 2004;20:99–108.

[18] Sheldon Weisgrau. Issues in rural health: access, hospitals, and reform Health Care Financal Review. 1995;17:1–14.

[19] O’Neal Wesley T, Sandesara Pratik B, Kelli Heval M, Venkatesh Sanjay, Soliman Elsayed Z. Urban-rural differences in mortality for atrial fibrillation hospitalizations in the United States Heart Rhythm. 2018;15:175–179.

[20] James Paul A, Li Pengxiang, Ward Marcia M. Myocardial infarction mortality in rural and urban hospitals: rethinking measures of quality of care Ann. Fam. Med.. 2007;5:105–111.

[21] Davidson Thomas, Mirza Farhaan, Baig Mirza M. Impact of socio-economic status and race on emergency hospital admission outcomes: Analysis from hospital admissions between 2001 and 2012 Health Serv. Manage. Res.. 2022;35:127–133.

